# Awareness and use of Tranexamic Acid in the management of Postpartum Haemorrhage among health care providers in Enugu, Nigeria

**DOI:** 10.1101/2023.06.02.23290525

**Authors:** Ogochukwu Onwujekwe, Adaobi Uchenna Mosanya, Kingsley Ekwuazi, Chukwuemeka Iyoke

**Author notes:** **Corresponding author:** Adaobi Uchenna Mosanya, Department of Clinical Pharmacy and Pharmacy Management, University of Nigeria, Nsukka, **Faculty of Pharmaceutical Sciences building, Chime Avenue, University of Nigeria, Nsukka, Enugu State, Nigeria. P.M.B. 410001**.

## Abstract

**Objectives:** This study was carried out to determine the knowledge, practice and barriers regarding the use of Tranexamic acid for the prevention and treatment of postpartum hemorrhage among health care providers in Enugu, Nigeria.

**Methods:** A cross sectional study was carried out among health professionals (Doctors, Pharmacists and Nurses) in two Nigerian tertiary teaching hospitals. One is Federal and the other is State. A total of 220 questionnaires were distributed, 207 were collected back (response rate: 94%) and analyzed using SPSS for inferential statistics with level of significance at p < 0.05.

**Results:** Only 23.7% of the respondents had good Knowledge (p<0.001). Good awareness of the recent WHO recommendation on the use of Tranexamic acid for Postpartum Haemorrhage was low (19.8%, <0.001). Majority of the respondents have neither prescribed nor dispensed it (30%; <0.001). Very few respondents use it for all cases of Postpartum Haemorrhage (16.4%; p<0.001). Barriers against its use include non-awareness of the latest WHO recommendation, preference for other utero-tonics and cost of the drug.

**Conclusions:** There was poor knowledge of Tranexamic acid, poor awareness of its recommendation and low use for Postpartum Haemorrhage among different cadres of health care providers.

**Synopsis:** There was poor knowledge of Tranexamic acid, poor awareness of its recommendation and low use for Postpartum Haemorrhage among different cadres of health care providers.

## Introduction

Maternal mortality is a major public health issue and maternal related causes accounted for 10.7 million deaths 1990 and 2015^1^. Globally an estimate of 303,000 maternal deaths occurred in 2015 mostly in the low resource settings, and most of them could have been prevented ^1^ Nigeria has one of the highest ratios of maternal mortality in the world accounting for approximately 58,000 maternal deaths i.e. 19% globally ^1^. One out of five causes of 70% of maternal mortality in Nigeria is hemorrhage ^2^ In April,2016, The World Maternal Antifibrinolytic trial (WOMAN) showed that showed Tranexamic acid when given early to women with Postpartum Haemorrhage could prevent one in three women from dying ^3^. As a consequence, the World Health Organization (WHO) published an updated recommendation on the use of Tranexamic acid (TXA) for Postpartum Haemorrhage treatment ^4^. Since then, several studies have been done to evaluate the safety and effectiveness (including cost-effectiveness) of Tranexamic acid for the prevention and treatment of postpartum hemorrhage, which may occur after vaginal as well as cesarean deliveries ^5–14^

Currently, there is no published study on the awareness of health care professionals regarding the use of Tranexamic acid for the prevention and management of Postpartum Haemorrhage in Nigeria. Therefore, this study aimed to determine the knowledge and practice of health care providers in Enugu Metropolis on the use of Tranexamic acid for the management of Postpartum Haemorrhage and the barriers to effective adoption and implementation of the new recommendation. The findings of the work is expected to have important implications for the delivery of high quality maternal care and prevention of maternal death through Postpartum Haemorrhage.

## METHODS

### Study design

The study was a cross sectional study and data were collected using a structured pre tested questionnaire for health providers involved in obstetrics and Gynecology in both hospitals. The respondents were medical doctors, nurses and pharmacists. A consecutive sampling method was used in administering the questionnaires.

### Study setting

The study was carried out in Enugu metropolis. Enugu is the capital city of Enugu State. There are two tertiary public hospitals and several private hospitals (three of which are faith based). The inhabitants of Enugu are mostly civil servants, although the city is increasingly acquiring commercial activities. The population is comprised of mostly people who are mainly Igbo by tribe and Christian by religion. The study was carried out in Enugu State which has two tertiary teaching hospitals with Obstetrics and Gynaecology units. Each of these centres records about 1000-3000 deliveries per annum. The study population comprised of healthcare providers (doctors, Pharmacists and nurses) working with the Obstetrics and Gynaecology units.

### Sampling method

A convenient sample of 220 questionnaires was administered to the respondents. A proportionate allocation was used to get the sample of respondents in the two hospitals based on the population of appropriate healthcare providers in both hospitals. Hence, 145 was allocated to the Federal teaching hospital while 75 was to the State teaching hospital.

### Data collection instrument

A pre-tested structured questionnaire was used to collect data. The questionnaire was divided into 5 sections, the first section was on the Social demographic characteristics, the second was on Awareness and knowledge of respondents on the use of Tranexamic acid for Postpartum Haemorrhage (PPH) treatment and prevention, the third section was on provider practices, the fourth section was on the use of Tranexamic acid for Postpartum Haemorrhage and the last section was on acceptability and barriers to use of Tranexamic acid for Postpartum Haemorrhage.

### Data collection and analysis

Copies of the questionnaire were distributed to the respondents by the researcher after a brief introduction of the research work, the purpose of the study and obtaining verbal consent from the prospective participants. Data collection began towards the end of 2019. However, it was interrupted by COVID-19 pandemic and the subsequent lockdown. It was finally concluded in 2021. Data was analyzed using Statistical Package for Social Sciences (SPSS) version 25 and Microsoft Excel 2019. The results from the data analysis were presented using descriptive and inferential statistics. Frequencies and percentages were used to summarize the items on the various sections of the questionnaire.

### Outcome measurement

An overall knowledge/Awareness, Practice and Use was determined by scoring each correct response made by the respondents and summed them up to get the total score. The score was categorized as good if (> 50%) and poor (<50%). The inferential statistics –the Chi square Test of significance was used to assess the relationship between knowledge of Tranexamic acid and professions, institutions as well as the relationship between the use of Tranexamic acid and professions, institutions. The relationship was termed significant if p-value is less than .05 (p<.05).

### Ethical Consideration

Ethical clearance was obtained from the ethical committee of the University of Nigeria Teaching Hospital and informed consent was obtained from all respondents before completing the questionnaire and those that declined were left out.

## RESULTS

A total of 207 questionnaires were returned and analyzed. This comprised of seventy (70) respondents (response rate: 93.33%) from the Enugu state university teaching hospital and one hundred and thirty seven (137) respondents (response rate: 94.48%) from the University of Nigeria Teaching Hospital Ituku Ozalla, Enugu State.

### Socio Demographic Characteristics

Approximately half (51.7%) of the respondents were medical doctors, 46.4 % were within the ages 30-39 years, while 57% were females (Table 1) Regarding their cadre, the doctors were mainly house officers (48.6%) while the pharmacists were mainly interns (29.5%) and the nurses were majorly staff nurses/midwives (56.4%). A greater percentage of the participants had bachelor’s degree as their highest education level (67.6%) and those with less than 5 years of practice accounted for 39.6%.

**Table 1:** Demographic Characteristics of the Health Professionals.

|  |  | Frequency | Percent |
| --- | --- | --- | --- |
| **Age** | <30 | 66 | 31.9 |
|  | 30-39 | 96 | 46.4 |
|  | 40-49 | 36 | 17.4 |
|  | 50-59 | 8 | 3.9 |
|  | ≥ 60 | 1 | 0.5 |
| **Sex** | Male | 89 | 43.0 |
|  | Female | 118 | 57.0 |
| **Profession** | Medical doctor | 107 | 51.7 |
|  | Pharmacist | 61 | 29.5 |
|  | Nurse/midwife | 39 | 18.8 |
| **Medical Doctors' cadre**<br>**(n = 107)** | House officer | 52 | 48.6 |
|  | Registrar | 44 | 41.1 |
|  | Senior registrar | 5 | 4.7 |
|  | Consultant | 6 | 5.6 |
| **Cadre of Pharmacists**<br>**(n = 61)** | Intern | 18 | 29.5 |
|  | Senior pharmacist | 12 | 19.7 |
|  | Principal pharmacist | 12 | 19.7 |
|  | Chief pharmacist | 8 | 13.1 |
|  | Assistant director | 9 | 14.8 |
|  | Deputy director | 2 | 3.3 |
| **Nurses' cadre**<br>**(n = 39)** | Intern | 2 | 5.1 |
|  | Staff nurse/midwife | 22 | 56.4 |
|  | Senior nursing officer | 8 | 20.5 |
|  | PNO | 1 | 2.6 |
|  | Matron | 3 | 7.7 |
|  | CNO | 1 | 2.6 |
|  | Senior matron | 2 | 5.1 |
| **Highest level of education** | Diploma | 13 | 6.3 |
|  | Bachelors/1st degree | 140 | 67.6 |
|  | Master's degree | 24 | 11.6 |
|  | Professional fellowship | 22 | 10.6 |
|  | No response | 8 | 3.9 |
| **Years of practice in profession** | < 5 | 82 | 39.6 |
|  | 5-10 | 70 | 33.8 |
|  | 11-15 | 27 | 13.0 |
|  | 16-20 | 13 | 6.3 |
|  | 21-25 | 3 | 1.4 |
|  | 26-30 | 3 | 1.4 |
|  | > 30 | 3 | 1.4 |
|  | No response | 6 | 2.9 |

### Knowledge on the Use of Tranexamic Acid for postpartum hemorrhage

A good proportion of the respondents (85.5%) knew that post-partum haemorrhage associated deaths could be avoided by the use of prophylactic uterotonics during the third stage of labor (Table 2). However, very few (14%) knew that the conclusion of the Guideline Development Group meeting on the prevention and treatment of post-partum haemorrhage in March 2012 was that there was no direct evidence of the effectiveness and safety of tranexamic acid for post-partum haemorrhage. Knowledge of tranexamic acid as a synthetic lysine analog trans-4 aminoethylcyclo hexanecarboxylic acid was about average (50.2%); those who knew that the mechanism of action of tranexamic acid was by binding to plasminogen molecules, thereby blocking the binding of lysine to them were almost average (45.9%). In summary, the proportion of the respondents with good knowledge regarding the use of Tranexamic acid for Postpartum Haemorrhage was very low (Table 3)

**Table 2:** Knowledge about use of tranexamic acid for postpartum haemorrhage.

|  | Yes | No | Don't know | Chi-Square | P-value |
| --- | --- | --- | --- | --- | --- |
| Postpartum hemorrhage is defined as blood loss of 1000ml or more within 24 hours after birth | 157(75.8) | 40(19.3)<sup>a</sup> | 10(4.8) | 174.870 | <.001 |
| Majority of PPH associated deaths could be avoided by the use of prophylactic uterotonics during the third stage of labour | <sup>a</sup>177(85.5) | 5(2.4) | 25(12.1) | 256.464 | <.001 |
| The Guideline Development Group meeting on the prevention and treatment of PPH in March 2012 concluded there was no direct evidence of the effectiveness and safety of TXA for PPH | 29(14.0)<sup>a</sup> | 43(20.8) | 135(65.2) | 96.116 | <.001 |
| TXA is a synthetic lysine analog trans-4 aminoethylcyclohexanecarboxylic acid | <sup>a</sup>104(50.2) | 3(1.4) | 100(48.3) | 94.812 | <.001 |
| The mechanism of action of TXA is by binding to plasminogen and thereby blocking activation to plasmin leading to inhibition of fibrinolysis and fibrinogenolysis | 95(45.9)<sup>a</sup> | 32(15.5) | 80(38.6) | 31.391 | <.001 |
| The April 2017 world maternal anti-fibrinolytic (WOMAN) trial supersedes the March 2012 trial | 52(25.1)<sup>a</sup> | 3(1.4) | 152(73.4) | 167.159 | <.001 |
| The woman trial for PPH concluded on oral and intravenous administration of TXA for PPH | 83(40.1) | 11(5.3)<sup>a</sup> | 113(54.6) | 79.652 | <.001 |
| PPH is a leading cause of maternal mortality with about one quarter of all maternal deaths associated with it in low income countries | <sup>a</sup>184(88.9) | 3(1.4) | 20(9.7) | 289.594 | <.001 |

<sup>a</sup>**indicates the right responses- correct knowledge or use**

**Table 3:** The categorized knowledge score about use of TXA for PPH.

|  |  | Frequency | Percent | Chi-Square | P-Value |
| --- | --- | --- | --- | --- | --- |
| Good | (Knowledge score > 50%) | 49 | 23.7 | 57.40 | <.001 |
| Poor | (Knowledge score ≤ 50%) | 158 | 76.3 |  |  |

### Provider Practices of Tranexamic acid for postpartum hemorrhage

Some of the providers (28%) perceived that tranexamic acid should be used in all cases of post-partum haemorrhage regardless of if bleeding is due to genital tract trauma or other causes (Table 4). However, majority believed that tranexamic acid should be recognized as a life-saving intervention and made readily available for the management of Postpartum Haemorrhage in settings where obstetric care is provided (67.1%).

**Table 4:** Provider’s use of TXA for PPH.

|  | Yes | No | Don't know | Chi-Square | P-value |
| --- | --- | --- | --- | --- | --- |
| TXA should be used in all cases of PPH regardless of if bleeding is due to genital tract trauma or other causes | 58(28.0) <sup>a</sup> | 86(41.5) | 63(30.4) | 6.46 | .039 |
| Have you recommended/dispensed TXA for the management or prevention of PPH? | 62(30.0) <sup>a</sup> | 129(62.3) | 16(7.7) | 93.59 | <.001 |
| Do you use TXA in all cases of PPH regardless of if bleeding is due to genital tract trauma or other causes? | 34(16.4) <sup>a</sup> | 160(77.3) | 13(6.3) | 183.22 | <.001 |
| Do you recognize TXA as a life-saving intervention for managing PPH in obstetrics practice? | <sup>a</sup>155(74.9) | 38(18.4) | 14(6.8) | 164.96 | <.001 |
| Have you used TXA for other cases of bleeding or haemorrhage other than for PPH? | 74(35.7) <sup>a</sup> | 102(49.3) | 31(15.0) | 37.07 | <.001 |

<sup>a</sup>**indicates the right responses- correct knowledge or use**

### Attitude on the use of Tranexamic acid for postpartum hemorrhage

Approximately a quarter of the respondents (21.7%) indicated the recommended dosage regimen of 1g (100mg/ml) IV over 10 min (21.7%) and below average number (42.5%) indicated that the effect of timing in administration of Tranexamic acid was very significant (Table 5). Indication of 1---3 hours after child birth as the best range of time to administer Tranexamic acid for optimal result was very poor (14.5%) while the indication that the use of tranexamic acid in women with contraindication to antifibrinolytic therapy not being a part of current routine care for Postpartum Haemorrhage treatment was about average (52.2%). Majority however perceived that Tranexamic acid should be considered as part of the standard postpartum haemorrhage treatment package (71.5%).

**Table 5:** Use of tranexamic acid for postpartum haemorrhage.

|  | Frequency | Percent | Chi-Square | P-Value |
| --- | --- | --- | --- | --- |
| **Do you use/dispense tranexamic acid for the management of post-partum haemorrhage in your facility?** |  |  | 2.74 | .098 |
| - Yes<sup>a</sup> | 85 | 41.1 |  |  |
| - No | 108 | 52.2 |  |  |
| - No response | 14 | 6.8 |  |  |
| **Recommended dosage regimen used for tranexamic acid in PPH treatment?** |  |  | 239.06 | <.001 |
| - 2g (200mg/ml) IV over 10 min | 19 | 9.2 |  |  |
| - 3g (300mg/ml) IV over 20 min | 14 | 6.8 |  |  |
| - 1g (100mg/ml) IV over 10 min<sup>a</sup> | 45 | 21.7 |  |  |
| - 4g (400mg/ml) IV over 10 min | 3 | 1.4 |  |  |
| - Don't know | 126 | 55.6 |  |  |
| **What is the effect of timing in the administration of tranexamic acid for post-partum haemorrhage** |  |  | 71.51 | <.001 |
| - Not significant | 4 | 1.9 |  |  |
| - Very significant<sup>a</sup> | 88 | 42.5 |  |  |
| - Not sure | 53 | 25.6 |  |  |
| - Don't know | 62 | 28.5 |  |  |
| **What is the best range of time to administer tranexamic acid for optimal result** |  |  | 256.10 | <.001 |
| - Immediately after child birth | 58 | 28.0 |  |  |
| - 1-3 hours after child birth<sup>a</sup> | 30 | 14.5 |  |  |
| - 4-6 hours after child birth | 6 | 2.9 |  |  |
| - 7-10 hours after child birth | 2 | 1.0 |  |  |
| - > 10 hours after child birth | 3 | 1.4 |  |  |
| - Don't know | 108 | 52.2 |  |  |
| **Which of these is not a part of the current routine care for PPH treatment?** |  |  | 138.34 | <.001 |
| - Fluid replacement | 20 | 9.7 |  |  |
| - Surgical interventions | 24 | 11.6 |  |  |
| - Use of tranexamic acid in women with contraindication to antifibrinolytic therapy<sup>a</sup> | 108 | 52.2 |  |  |
| - Monitoring vital signs | 36 | 17.4 |  |  |
| - No response | 19 | 9.2 |  |  |
| **Tranexamic acid should be considered as part of the standard PPH treatment package?** |  |  | 57.87 | <.001 |
| - Strongly agree<sup>a</sup> | 73 | 35.3 |  |  |
| - Agree<sup>a</sup> | 75 | 36.2 |  |  |
| - Disagree | 7 | 3.4 |  |  |
| - Don't know | 52 | 25.1 |  |  |

<sup>a</sup>indicates the right responses- correct knowledge or use

### Barriers against use of Tranexamic acid for postpartum hemorrhage

There were various barriers identified as constraints to the use of Tranexamic acid by healthcare workers to manage Postpartum Haemorrhage. The most common barrier was ignorance about the latest recommendation by the WHO on the use of Tranexamic acid. The next most common barrier was preference for substitute drugs for the condition (Figure 1).

### Relationship profession, institution, knowledge of TXA for postpartum hemorrhage

There was no significant relationship observed between profession and Institution with knowledge of the use of Tranexamic acid for postpartum hemorrhage (Table 6) Proportion of those with good knowledge between the different professions was comparable: medical doctor (22.4%), pharmacist (27.9%) and nurse/midwife (20.5%); and likewise between institutions: UNTH (25.5%) and ESUTH (20.0%).

**Table 6:** Relationship between Profession, Institution and Knowledge of TXA for PPH.

|  |  | Knowledge on Use of TXA for PPH |  |  | Chi-Square | p-value |
| --- | --- | --- | --- | --- | --- | --- |
|  |  | Good (n = 49) | Poor (n = 158) | Total (n = 207) |  |  |
| Profession | Medical doctor | 24(22.4) | 83(77.6) | 107 | .901 | .637 |
|  | Pharmacist | 17(27.9) | 44(72.1) | 61 |  |  |
|  | Nurse/midwife | 8(20.5) | 31(79.5) | 39 |  |  |
| Institution | UNTH | 35(25.5) | 102(74.5) | 137 | .789 | .374 |
|  | ESUTH | 14(20.0) | 56(80.0) | 70 |  |  |

### Relationship of profession, institution, use of TXA for postpartum hemorrhage

The use of Tranexamic acid for postpartum hemorrhage had significant relationship with both profession (p < .001) and institution (p < .001). Specifically, for profession, use of Tranexamic acid for postpartum hemorrhage was highest among pharmacists (61.8%) followed by medical doctors (44.0%) and the least was nurses/midwives (18.4%). More than half of the respondents from the UNTH (59.7%) used Tranexamic acid for postpartum hemorrhage unlike their counterparts in ESUTH (Table 7). In addition, there was a significant relationship between knowledge about the use of Tranexamic acid for postpartum hemorrhage and the use of it (p < .001). Those with good knowledge on the use (66.7%) were associated with the actual use far more than those with poor knowledge (37.2%). More specifically, the odds of use of Tranexamic acid for postpartum hemorrhage was 3.4 times [95% C.I-1.67-6.84] higher among those with good knowledge than those with poor knowledge.

**Table 7:** Relationship between Profession, Institution and Use of TXA for PPH.

|  |  | Use of TXA for Postpartum hemorrhage |  |  | Chi-Square | p-value |
| --- | --- | --- | --- | --- | --- | --- |
|  |  | Yes (n = 85) | No (n = 108) | Total (n = 207) |  |  |
| Profession | Medical doctor | 44(44.0) | 56(56.0) | 100 | 17.174 | < .001 |
|  | Pharmacist | 34(61.8) | 21(38.2) | 55 |  |  |
|  | Nurse/midwife | 7(18.4) | 31(81.6) | 38 |  |  |
| Institution | UNTH | 74(59.7) | 50(40.3) | 124 | 34.407 | < .001 |
|  | ESUTH | 11(15.9) | 58(84.1) | 69 |  |  |

## DISCUSSION

This study has the strength of being the first because there are no studies in published literature assessing the knowledge and use of Tranexamic acid for postpartum hemorrhage among health care providers. Majority of the surveyed health providers failed the definition of postpartum hemorrhage which is generally defined as blood loss of 500 ml and above after delivery through the vagina or blood loss of 1000 ml and above after delivery through cesarean section ^15^. Approximately, half of the respondents in this current study understood the mechanism of action of Tranexamic acid. Medical or health care professionals in gynecology should have the knowledge of the mechanism of action of Tranexamic acid. The mechanism of action of Tranexamic acid as explained by Cai and colleagues is brought about through blocking the binding site for lysine on plasminogen molecules thereby preventing the association of plasminogen with already formed plasmin and fibrin thus stopping rapid fibrinolysis of the formed fibrin mesh ^16^

There was poor use of Tranexamic acid and even among providers that use it, majority didn’t know the recommended dose. An average proportion of the respondents were not sure of the appropriate dose of Tranexamic acid for the management of postpartum hemorrhage. In fact a minimal percentage of them got the correct dose which was a loading dose of 1 g (100 mg/mL) intravenously given at a rate of 1 mL/min ^3^ Tranexamic acid is one of the evidence-based approaches to reducing the incidence of adverse maternal outcomes associated with PPH. Other equally important approaches are prompt blood transfusion as well as active management of the third stage of labor ^17^. The proportion of the respondents who agreed that timing plays a significant role in the administration of Tranexamic acid were almost the same proportion as those who thought that immediate administration and 1-3 hours after birth put together. Those who thought that the administration of Tranexamic acid immediately after child birth was the best time range of Tranexamic acid for optimum outcome were much more that those who got the correct time range which was between 1 to 3 hours after child birth. In reality, the time range with evident advantage was between 1-3 hours. In fact it was observed that this timely administration significantly reduced the risk of mortality due to bleeding as well as the reliance on laparotomy for bleeding control ^3^. The poor knowledge and use observed in this current study could be attributed to not being updated on current guidelines and also lack of advocacy and training of health professional. Importance of training can never be over emphasized regarding emergency obstetrics response. There is evidence that trainings aimed to improve clinical knowledge and skills with respect to the risk factors of postpartum hemorrhage prevents maternal morbidity and mortality^17^.

Few respondents have recommended, dispensed or administered and actually used it in practice despite recognizing it as a lifesaving intervention that should be made available for the management of Tranexamic acid in Obstetrics practice. They were more likely to use it for other cases of bleeding than postpartum hemorrhage. It shows that there is still room for greater awareness to be created for the use of Tranexamic acid in postpartum hemorrhage management. Possible avenues include clinical meetings and continuous professional development sessions. This suboptimal practice could also be as a result of not being updated on the most current guideline. Perhaps other reasons could be related to the barriers against its use as observed in this current study. Some of the perceived barriers were ignorance of the latest recommendation, preference to other uterotonics, high cost of drug, and fear of thromboembolic event and drug scarcity.

One of the barriers identified in this study against the optimal use of Tranexamic acid for the management of postpartum hemorrhage was high cost of the former. It is important to highlight here some of the outcomes from cost-effectiveness studies done on the use of Tranexamic acid for postpartum hemorrhage. Li Bernadette and colleagues calculated the incremental cost-effectiveness ratios (ICER) of treating postpartum hemorrhage within 3 hours of giving birth with or without Tranexamic acid. Results generated were optimistic. Despite the possibility of increasing the relative risk of death due to bleeding while on Tranexamic acid to the upper bound of 0.91, the ICER was $ 692 per Quality-adjusted life-Years (QALY). In addition, if the discount rate was applied as high as 10%, the ICER ($ 507 per QALY) was still within the threshold range of Nigeria ($ 446 - $ 2880 per QALY) i.e. within the cost-effective range. In summary, at the lower threshold range ($ 446 per QALY) the probability that Tranexamic acid is cost-effective is 93% ^18^ This is comparable with the results from a more recent study in a developed country ^6^

## CONCLUSION

The findings show that there is poor knowledge about the use of WHO recommended Tranexamic acid for postpartum hemorrhage among different cadres of healthcare workers. The uptake of the recommendation among health workers was rather low and seriously affecting its use. Most health providers have a poor awareness of the recommendation which led to a poor practice and use.

## Data Availability

All data produced in the present study are available upon reasonable request to the authors

## RECOMMENDATIONS

A major limitation of this study was the cross sectional design adopted. Therefore there is need for prospective studies which would provide a more robust evidence on the cost effectiveness of Tranexamic acid for postpartum hemorrhage. This will provide basis for the advocacy by policy makers targeting professionals and updating them on current best practices and also alleviating their fears.

The authors declare no conflict of interest

There was no external funding for this study

## Authors’ contribution

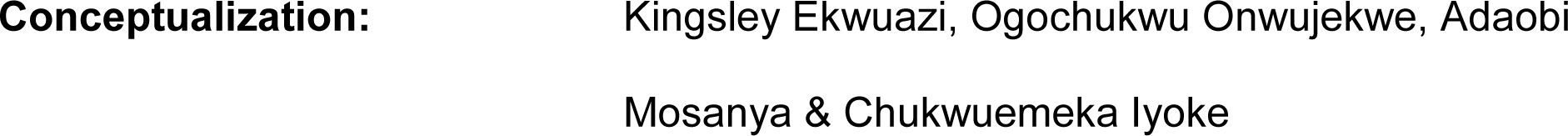
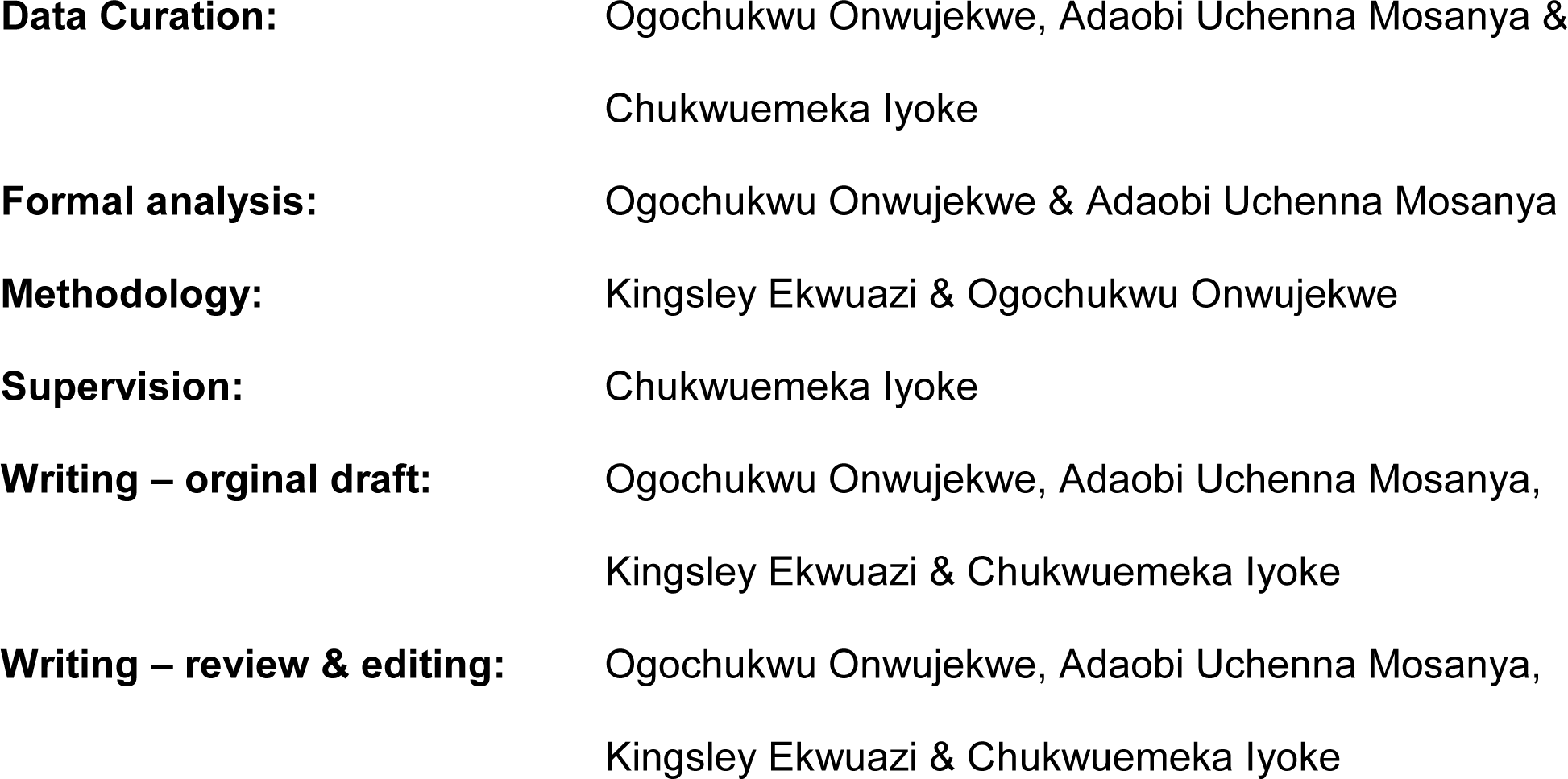

## REFERENCES

1. Say L, Chou D, Gemmill A, et al. Global causes of maternal death: A WHO systematic analysis. Lancet Glob. Health. 2014;2(6). doi:10.1016/S2214-109X(14)70227-X

2. Piane GM. Maternal Mortality in Nigeria: A Literature Review. World Med. Health Policy. 2019;11(1):83–94. doi:10.1002/wmh3.291

3. Shakur H, Roberts I, Fawole B, et al. Effect of early tranexamic acid administration on mortality, hysterectomy, and other morbidities in women with post-partum haemorrhage (WOMAN): an international, randomised, double-blind, placebo-controlled trial. Lancet. 2017;389(10084):2105–2116. doi:10.1016/S0140-6736(17)30638-4

4. World Health Organization. Updated WHO Recommendation on Tranexamic Acid for the Treatment of Postpartum Haemorrhage: Highlights and Key Messages from the World Health Organization’s 2017 Global Recommendation. 2017;(October):5.

5. Sudhof LS, Shainker SA, Einerson BD. Tranexamic acid in the routine treatment of postpartum hemorrhage in the United States: a cost-effectiveness analysis. Am. J. Obstet. Gynecol. 2019;221(3):275.e1-275.e12. doi:10.1016/j.ajog.2019.06.030

6. Howard DC, Jones AE, Skeith A, Lai J, D’Souza R, Caughey AB. Tranexamic acid for the treatment of postpartum hemorrhage: a cost-effectiveness analysis. Am. J. Obstet. Gynecol MFM. 2022;4(3):100588. doi:10.1016/j.ajogmf.2022.100588

7. Modir H, Moshiri E, Naseri N, Faraji F, Almasi-Hashiani A. A randomized parallel design trial of the efficacy and safety of tranexamic acid, dexmedetomidine and nitroglycerin in controlling intraoperative bleeding and improving surgical field quality during septorhinoplasty under general anesthesia. Med. Gas Res. 2021;11(4):131–137. doi:10.4103/2045-9912.318857

8. Wong J, George RB, Hanley CM, Saliba C, Yee DA, Jerath A. Tranexamic acid: current use in obstetrics, major orthopedic, and trauma surgery. Can J Anaesth 2021;68(6):894–917. doi:10.1007/s12630-021-01967-7

9. Franchini M, Mengoli C, Cruciani M, et al. Safety and efficacy of tranexamic acid for prevention of obstetric haemorrhage: An updated systematic review and meta-analysis. Blood Transfus. 2018;16(4):329–337. doi:10.2450/2018.0026-18

10. Eyeberu A, Getachew T, Amare G, et al. Use of tranexamic acid in decreasing blood loss during and after delivery among women in Africa: a systematic review and meta-analysis. Arch. Gynecol. Obstet.. 2022;(0123456789). doi:10.1007/s00404-022-06845-1

11. Wang K, Santiago R. Tranexamic acid – A narrative review for the emergency medicine clinician. Am. J. Emerg. Med. 2022;56:33–44. doi:10.1016/j.ajem.2022.03.027

12. Van Houwe M, Roofthooft E, Van de Velde M. The role of tranexamic acid in obstetric hemorrhage: a narrative review. Acta Anaesthesiol. Belg. 2022;73(2):103–108. doi:10.56126/73.2.12

13. Bouthors AS, Gilliot S, Sentilhes L, et al. The role of tranexamic acid in the management of postpartum haemorrhage. Best Pract. Res. Clin. Anaesthesiol. 2022;36:411–426. doi:10.1016/j.bpa.2022.08.004

14. Bellos I, Pergialiotis V. Tranexamic acid for the prevention of postpartum hemorrhage in women undergoing cesarean delivery: an updated meta-analysis. Am. J. Obstet. Gynecol. 2022;226(4):510–523.e22. doi:10.1016/j.ajog.2021.09.025

15. Smith JR. Medscape. Postpartum Hemorrhage. https://emedicine.medscape.com/article/275038-print. Published 2022. Accessed January 16, 2023.

16. Cai J, Ribkoff J, Olson S, et al. The many roles of tranexamic acid: An overview of the clinical indications for TXA in medical and surgical patients. Eur. J. Haematol. 2020;104(2):79–87. doi:10.1111/ejh.13348

17. Cooper N, O’Brien S, Siassakos D. Training health workers to prevent and manage post-partum haemorrhage (PPH). Best Pract Res Clin Obstet Gynaecol 2019;61(November 2019):121–129. doi:10.1016/j.bpobgyn.2019.05.008

18. Li B, Miners A, Shakur H, Roberts I. Tranexamic acid for treatment of women with post-partum haemorrhage in Nigeria and Pakistan: a cost-effectiveness analysis of data from the WOMAN trial. Lancet Glob. Health 2018;6(2):e222–e228. doi:10.1016/S2214-109X(17)30467-9

